# Persistence of functional memory B cells recognizing SARS-CoV-2 variants despite loss of specific IgG

**DOI:** 10.1101/2021.05.15.21257210

**Authors:** Stephan Winklmeier, Katharina Eisenhut, Damla Taskin, Heike Rübsamen, Celine Schneider, Peter Eichhorn, Oliver T. Keppler, Matthias Klein, Simone Mader, Tania Kümpfel, Edgar Meinl

## Abstract

While some COVID-19 patients maintain SARS-CoV-2-specific serum IgGs for more than 6 months post-infection, others, especially mild cases, eventually lose IgG levels. We aimed to assess the persistence of SARS-CoV-2-specific B cells in patients who have lost specific IgGs and analyzed the reactivity of the immunoglobulins produced by these B cells. Circulating IgG memory B cells specific for SARS-CoV-2 were detected in all 16 patients 1–8 months post-infection, and 11 participants had specific IgA B cells. Four patients lost specific serum IgG after 5–8 months but had SARS-CoV-2-specific-B-cell levels comparable to those of seropositive donors. Immunoglobulins produced after in vitro differentiation blocked receptor-binding domain (RBD) binding to the cellular receptor ACE-2, indicating neutralizing activity. Memory-B-cell-derived IgGs recognized the RBD of B.1.1.7 similarly to the wild-type, while reactivity to B.1.351 and P.1. decreased by 30% and 50%, respectively. Memory-B-cell differentiation into antibody-producing cells is a more sensitive method for detecting previous infection than measuring serum antibodies. Circulating SARS-CoV-2 IgG memory B cells persist, even in the absence of specific serum IgG; produce neutralizing antibodies; and show differential cross-reactivity to emerging variants of concern. These features of SARS-CoV-2-specific memory B cells will help to understand and promote long-term protection.

## Introduction

The development of adaptive immunity to SARS-CoV-2 may provide protection against re-infection and allows the identification of patients that have had a previous infection. Adaptive immunity to SARS-CoV-2 involves antibody (Ab)-producing cells, memory B cells, and several T cell subsets. Analysis of immune responses to different viruses, including other coronaviruses, has shown that the lifespans of the adaptive immune system components vary (1, 2).

Details of the kinetics of immune responses to SARS-CoV-2 are beginning to be uncovered (3-9). Antibody (Ab) responses peak at about 2–3 weeks after infection, at which point the Ab-levels decline (4, 10, 11). In most individuals, anti-SARS-CoV-2 serum Abs persist for more than 6 months after primary infection, but some patients rapidly lose their specific Abs, especially those that experienced a mild disease course (2, 4, 6, 10-13). It has been proposed that, in addition to serum antibody titers, the memory B cell pool should be evaluated to estimate humoral immunity as an indicator of immune protection (4, 14-16).

Initial Ab responses are made by short-lived plasmablasts that develop in extrafollicular sites (14, 17, 18), and the subsequent development of high-affinity and persistent Abs involves affinity maturation and the expansion of B cells in germinal centers (15, 19-22). Two types of B cells exit the germinal center: memory B cells and plasmablasts (14, 19). Many of these plasmablasts are short-lived and die within a few weeks, but some find survival niches in the bone-marrow and persist as long-lived plasma cells (23). The extent to which these long-lived plasma cells develop differs between different viruses and vaccines (24, 25). Different mechanisms regulate the survival of long-lived plasma cells (23, 26) and memory B cells (19).

Recently, SARS-CoV-2 variants of concern (VoCs) have emerged in the United Kingdom (B.1.1.7), South Africa (B.1.351), Brazil (P.1.) and elsewhere with multiple substitutions, some of which are in the RBD in the NTD and the receptor-binding motif (RBM) of the RBD (27, 28). These rapidly spreading VoCs are currently causing serious concerns regarding the increased frequency of re-infection, utility of convalescent plasma, and limited vaccine responses (29, 30).

In this study, we analyzed the persistence of IgA and IgG memory B cells specific for SARS-CoV-2 in COVID-19 patients. We specifically investigated donors who had lost circulating IgG to SARS-CoV-2 and analyzed whether they still harbored specific memory B cells in their blood. To study the patient B cells, we adopted a functional approach, converting blood-derived B cells into Ab-secreting cells in vitro (31-33). Having identified the SARS-CoV-2-specific memory B cells in the blood, we analyzed whether the secreted Abs have neutralizing activity and show cross-reactivity to the recently emerged SARS-CoV-2 VoCs B.1.1.7, B.1.351, and P.1. The findings of the study revealed functional properties of persisting memory B specific to SARS-CoV-2 and this could help to understand and promote protection.

## Results

### Persistence of IgG memory B cells specific for SARS-CoV-2 in the presence and absence of specific IgG

We analyzed, in parallel, the presence of memory B cells specific for SARS-CoV-2 in blood and specific IgG in serum (**Figure 1** shows our approach). Our study included 16 COVID-19 patients that had undergone a mild or asymptomatic disease course (**Table 1**), and pre-pandemic blood samples from six HC donors served as the control group. We detected B cells that could be developed into SARS-CoV-2-specific-IgG-secreting plasmablasts in the blood of all COVID-19 patients analyzed. The reactivity to SARS-CoV-2 of these in vitro differentiated plasmablasts and the patient sera from the same blood withdrawal was investigated (**Figure 2A**). Remarkably, the sera from three COVID-19 patients were negative in the ELISA, and a fourth was borderline. These four donors (HC=1, MS=2, SLE=1; #5, #11, #12, #15) had been seropositive 1–2 months after acute infection (**Table 1**) but had lost their specific IgG 5–8 months post-infection (HC=1, MS=2, SLE=1). Two of these four donors were under immunotherapeutic regimens at the time their blood was sampled for this study (**Table 1**).

**Figure 1.**
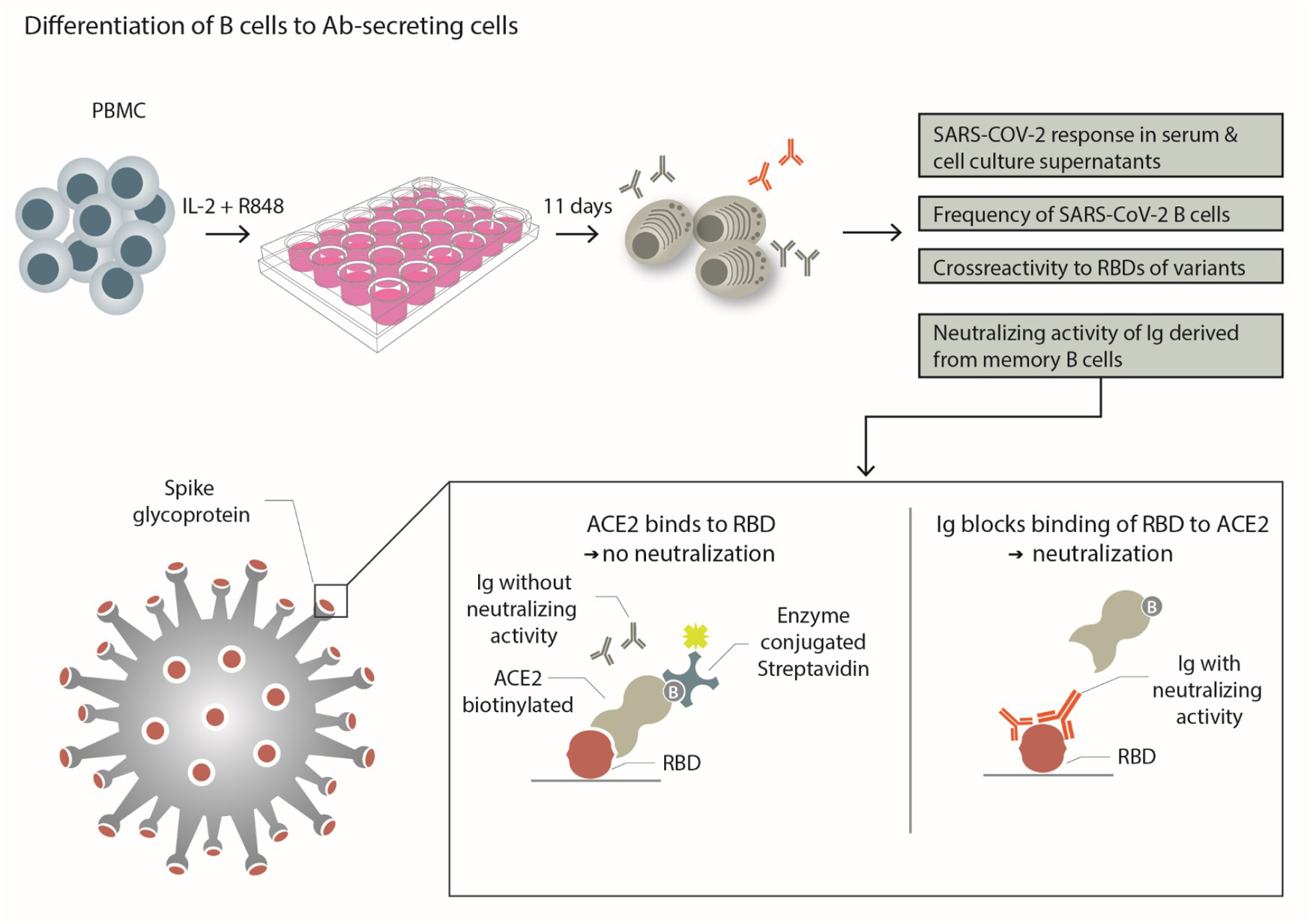
Experimental scheme. PBMCs from each donor were separated into individual wells and stimulated with the TLR7/8 agonist R848 and IL-2 to differentiate to them into Ab-secreting plasmablasts. This was used to compare the serum response to SARS-CoV-2 with that of specific Abs produced in vitro. The frequency of SARS-CoV-2-specific B cells that differentiated into Ab secreting cells was determined. The cross-reactivity to RBDs of emerging variants was tested. The ability of in vitro-produced Abs to block the binding of RBD to its receptor ACE-2 was determined as outlined.

**Table 1.**
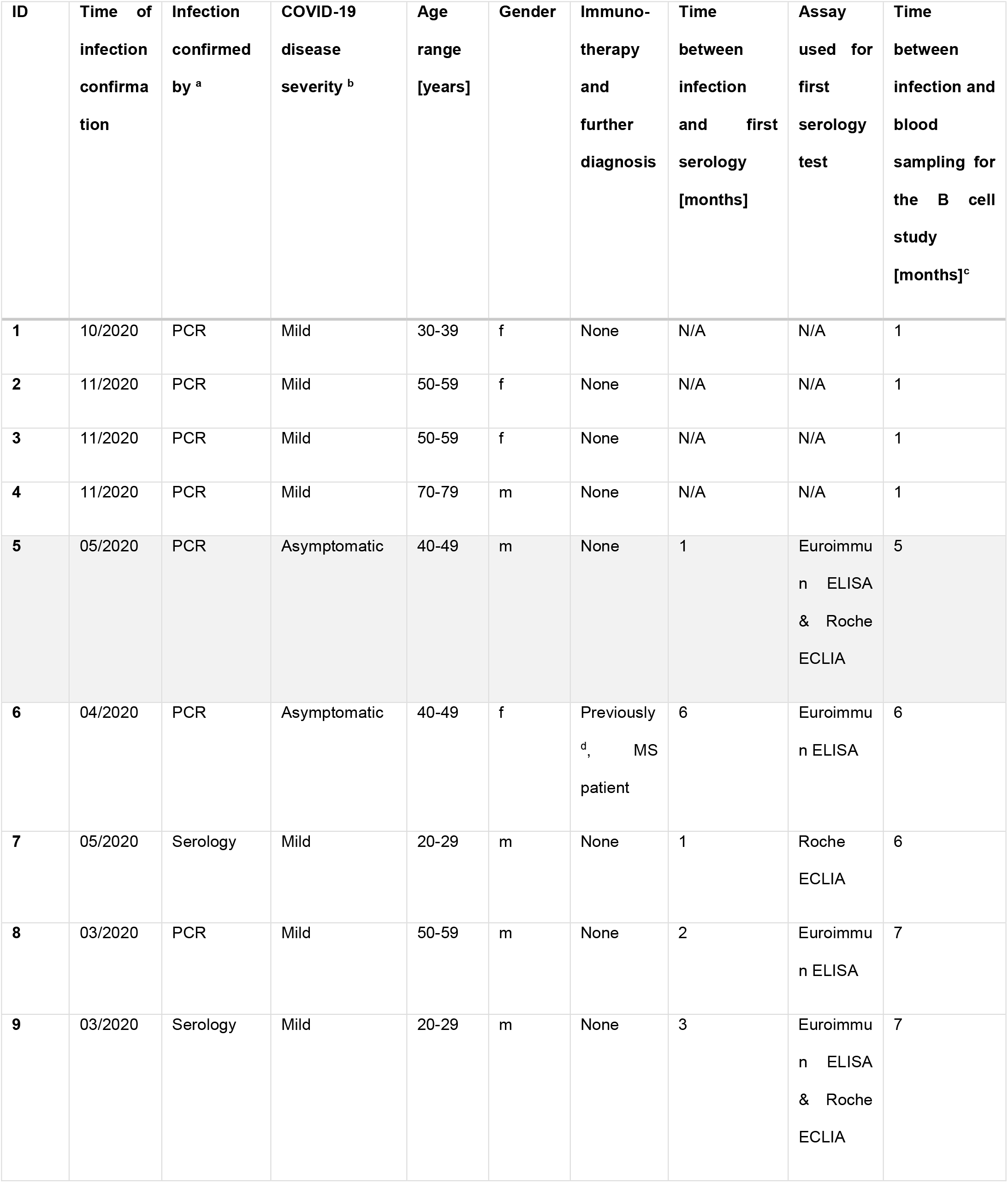
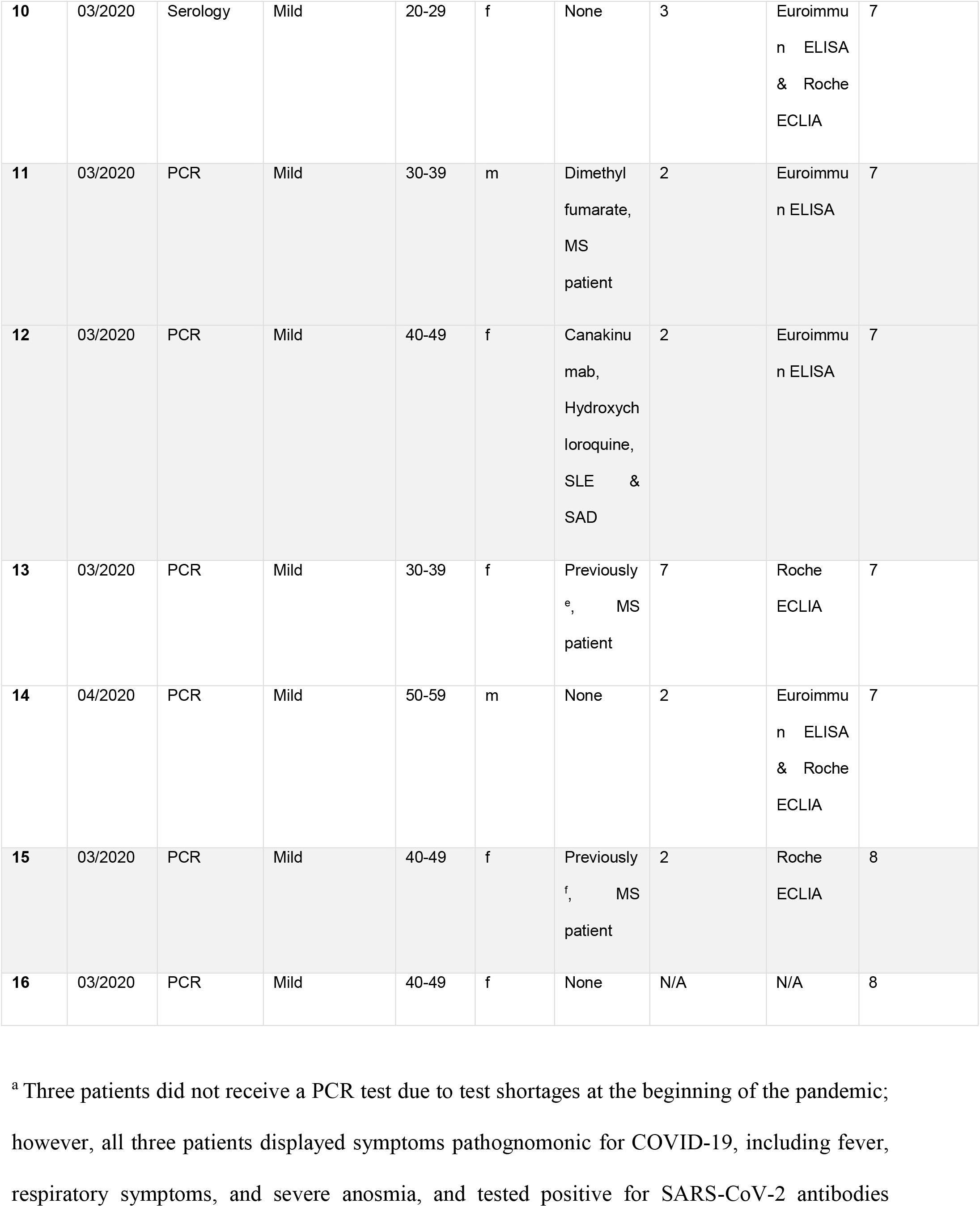
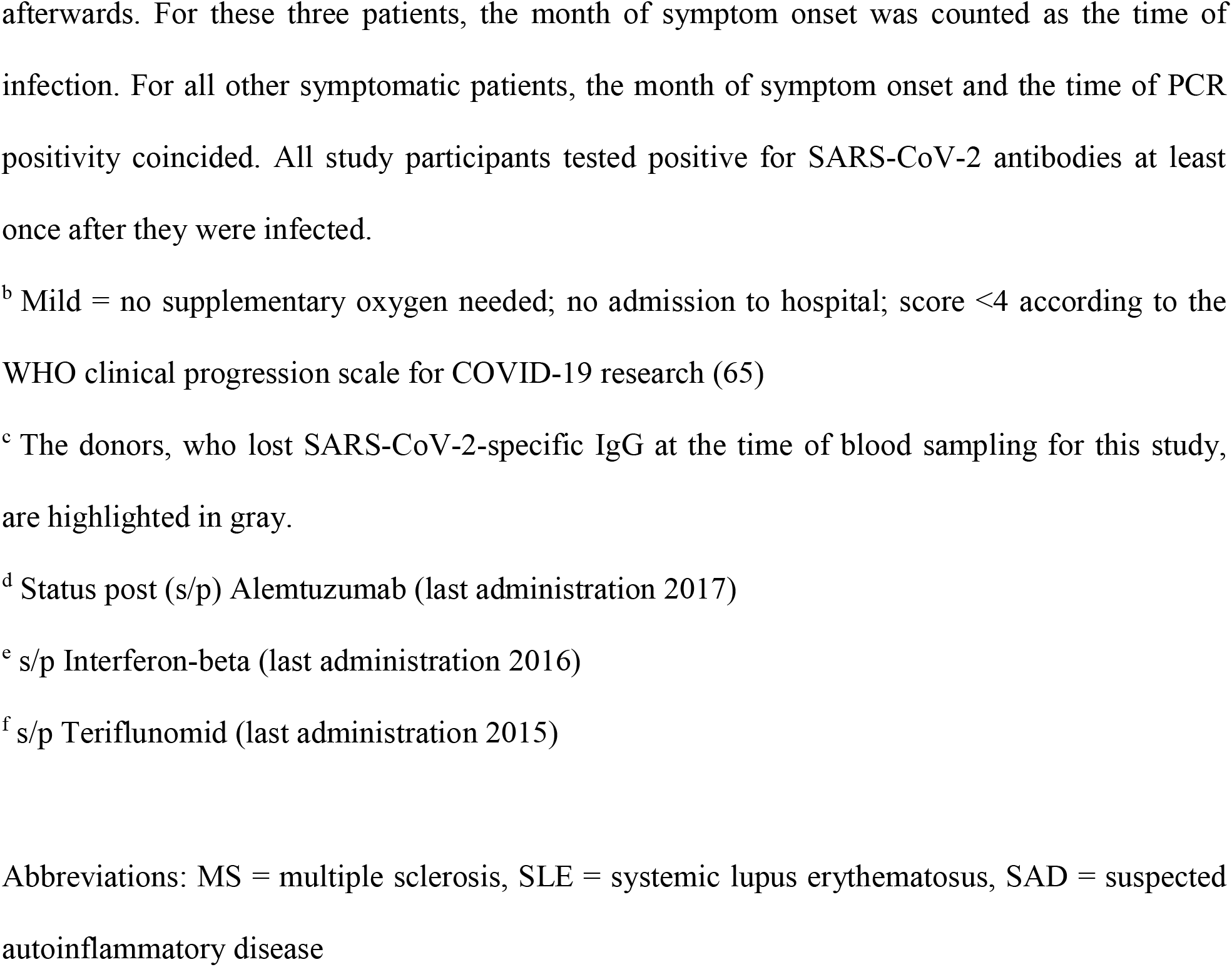
Characteristics of COVID-19 patients.

**Figure 2.**
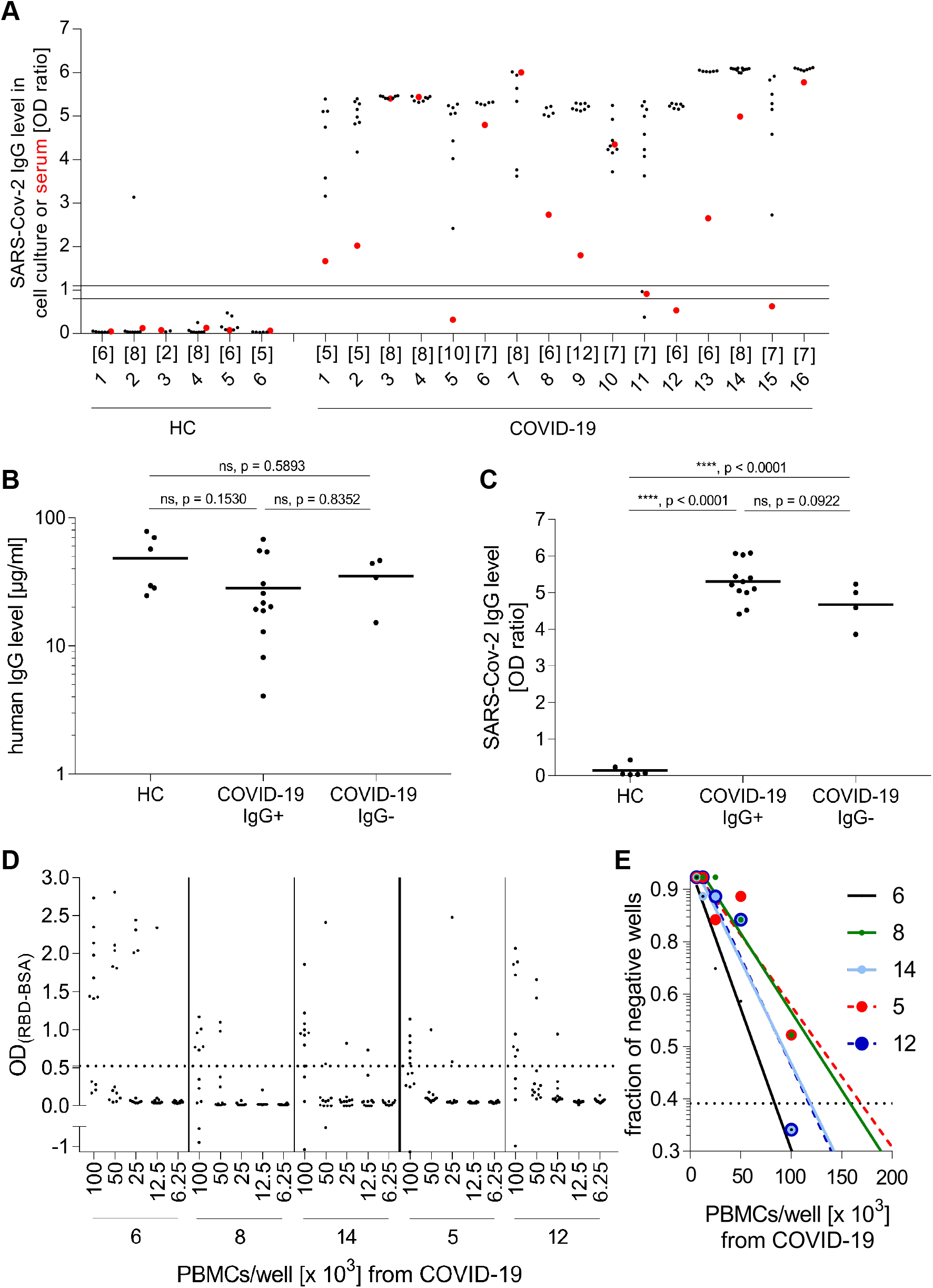
IgG production by differentiated B cells specific for SARS-CoV-2. (**A**) PBMCs from healthy controls (HC, left) and COVID-19-patients (right) were differentiated into Ab-secreting cells. The reactivity of IgG against S1 in cell culture supernatants was determined. Each black dot represents one stimulated well. The number of stimulated wells per donor is provided directly under the x-axis. The reactivity of the serum in the same ELISA is shown with a red dot. The area between the two horizontal lines were considered to represent the borderline zone of reactivity. COVID-patients who were serum-negative for SARS-CoV-2-specific IgG (#5, #11, #12, #15) were designated COVID-19-IgG^−^, and those who were positive were designated COVID-19-IgG^+^. (**B, C**) Each symbol represents the mean IgG level of all stimulated wells for one donor. Horizontal lines indicate the mean IgG levels of all donors in the respective groups. (**B**) IgG levels of cell culture supernatants were not significantly different between the groups (one-way ANOVA, Tukey’s multiple comparison test; HC=6, COVID-19-IgG^+^=12, COVID-19-IgG^−^=4) (**C**) Both the COVID-19-IgG^−^ and COVID-19-IgG^+^ subgroups produced more anti-S1 IgG (one-way ANOVA, Tukey’s multiple comparison test; HC=6, COVID-19-IgG^+^=12, COVID-19-IgG^−^=4) than the HC to a similar degree. (**D, E**) The raw data for the limiting dilution experiment are shown (**D**), and the calculation is displayed (**E**). Continuous lines are shown for donors who still had SARS-CoV-2 IgGs in their serum, and the dotted lines indicate donors who lost their specific serum Abs.

Based on these findings, we grouped the patients into those with (COVID19-IgG^+^) or without (COVID19-IgG^−^) serum IgG to SARS-CoV-2. We calculated the mean of the total IgG secreted into the cell culture supernatant and the SARS-CoV-2-specific IgG levels of the samples from each donor and compared the three groups (**Figure 2, B and C**). The COVID19-IgG^+^ and COVID19-IgG^−^ patients had significantly more SARS-CoV-2-specific B cells in their blood than HC donors (p < 0.0001), even though the amount of total secreted IgG was not significantly different between the groups. Remarkably, the levels of SARS-CoV-2-specific IgG produced in vitro were similar between the COVID19-IgG^−^ and COVID19-IgG^+^ subgroups **(Figure 2C)**.

The method of seeding PBMCs into individual wells and testing each well for the development of specific IgG yields a high sensitivity. We noted that all wells containing samples from 15 of the 16 COVID-19 patients were positive for SARS-CoV-2-specific IgG, and for one donor, 6 of the 8 wells were positive, which demonstrated the high frequency of SARS-CoV-2-specific B cells in these patients. We subsequently performed limiting dilution assays on PBMC from five COVID-19 patients (**Figure 2, D and E**). We calculated the frequency of specific B cells according to the Poisson distribution and the individual percentage of B cells within the PBMC (between 5% and 15%) and obtained thereby the following rates of B cells that gave rise to Abs to SARS-CoV-2: Patient #6 (seropositive): 1:13,000; patient #8 (seropositive): 1:11,000; patient #14 (seropositive): 1:20,000; patient #5 (seronegative): 1:22,000; patient #12 (seronegative) 1:7,000. Thus, in accordance with our calculation of SARS-CoV-2-reactivity in the bulk cultures (**Figure 2C**), our limiting dilution analysis indicated that the seronegative and seropositive COVID-19 patients had similar frequencies of circulating RBD-specific B cells. The frequencies observed lay within the reactivity ranges previously reported for measles virus and tetanus toxoid (32, 33). IgG memory B cells constitute about 15% of peripheral B cells, and only 30% to 40% of IgG memory B cells are capable of antibody production under these culture conditions (31). We found in one well of one HC sample B cells giving rise to SARS-CoV-2-recognizing Abs (**Figure 2A**); however, these Abs did not show neutralizing activity (see below).

We also analyzed the presence of anti-SARS-CoV-2 IgA in serum and of specific B cells in blood secreting IgA (**Supplemental Figure 1, A-C**). The COVID-19 patients had significantly more SARS-CoV-2-specific IgA B cells in their blood than the healthy controls (p = 0.022, Mann–Whitney U test). We noted that two COVID-19 patients had serum IgA, but no detectable specific IgA B cells. In healthy controls, SARS-CoV-2-specific IgA B cells were seen in 2 out of 6 donors and one was borderline, and no specific serum IgA was detected in 5 of the 6 healthy controls and 1 had borderline levels (**Supplemental Figure 1, A-C**). While all COVID-19 patients tested had specific IgG B cells in their blood, 11 patients had at least one well with specific IgA B cells. The difference between the occurrence of IgG- and IgA-positive B cells became clearer when we considered the number of positive wells. Specific IgG was detected in 115 out of 117 wells for COVID-19-patients, while specific IgA was detected in only 33 wells (p < 0.0001; p-values were determined using Fisher’s exact test). Therefore, IgA B cells specific for SARS-CoV-2 were detected in the blood of the COVID-19 patients but were less abundant than specific IgG B cells.

### Neutralizing activity of immunoglobulins derived from memory B cells

To analyze whether circulating peripheral B cells specific for SARS-CoV-2 can give rise to SARS-CoV-2-neutralizing Abs, we used a surrogate assay to analyze secreted-Ab inhibition of RBD binding to the viral entry receptor ACE-2 (34) (**Figure 1**). The significantly higher neutralizing potency of the COVID-19-patient-derived Abs produced in vitro was evident when compared to the neutralizing activity of the HC-derived Abs (**Figure 3**; p < 0.0001, Mann-Whitney U test). Thus, the SARS-CoV-2-specific B cells from COVID-19-patients released substantial amounts of neutralizing Abs after differentiation into Ab-secreting cells.

**Figure 3.**
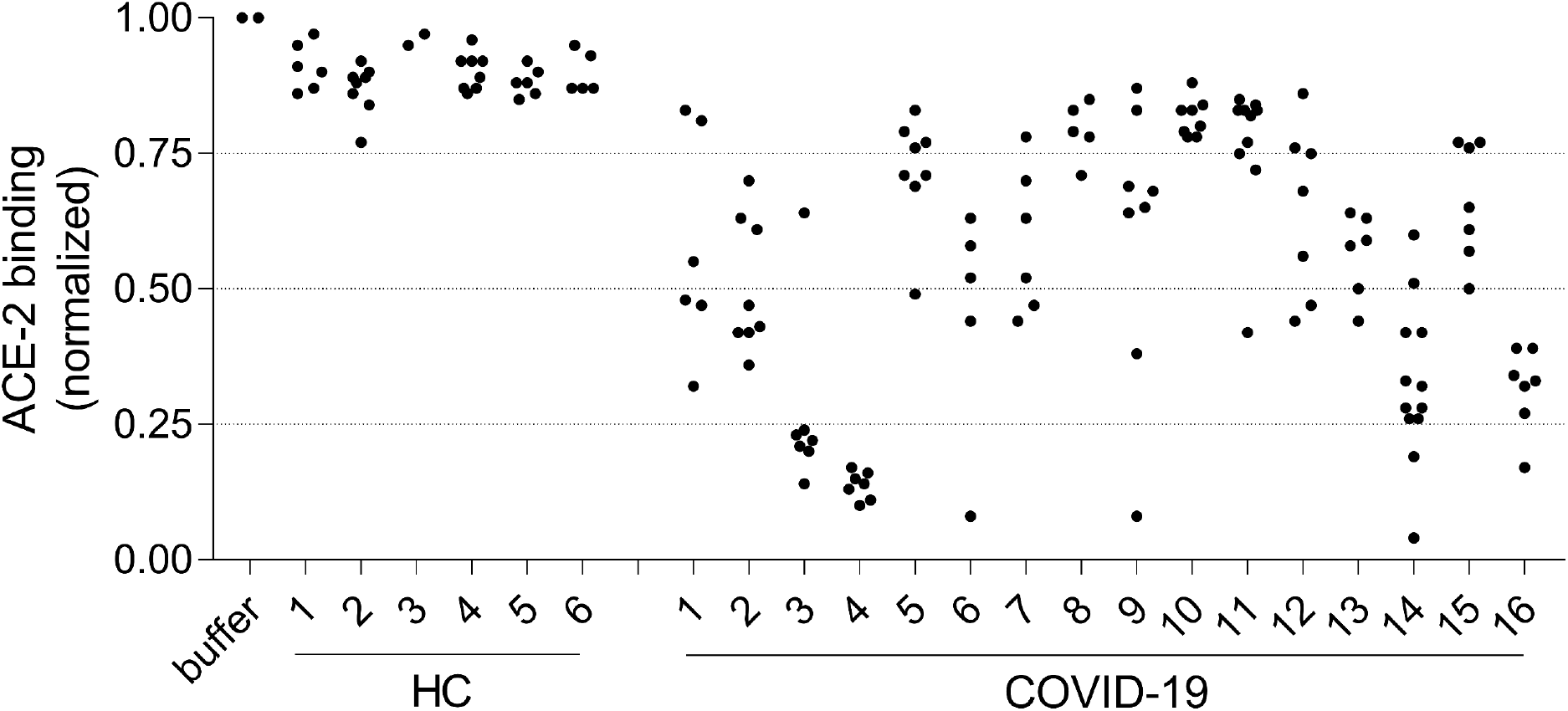
Neutralizing activity of Abs after differentiation of memory B cells. PBMCs from healthy controls (HC, left) and COVID-19 patients (right) were differentiated into Ab-secreting cells. The cell culture supernatants (each dot represents an individual well) were added to ELISA plates coated with the RBD. Biotinylated ACE-2 was then added, and its binding was detected with streptavidin–horseradish peroxidase. For calibration, the binding of biotinylated ACE-2 to RBD in the presence of buffer was set as 1. Then, the mean OD of the wells of each donor was calculated to compare the Abs binding to ACE-2 from COVID-19 patients with those from HCs. The Abs from COVID-19 patients reduced ACE-2 binding (p < 0.0001; Mann-Whitney U; HC=6, COVID-19=16).

### Cross-reactivity of B cells to variants of concern

We analyzed the cross-reactivity of SARS-CoV-2-specific memory B cells (**Figure 4A**) against the RBD of three major VoCs in current circulation: B.1.1.7, B.1.351, and P.1 (28). When we examined the memory B cells from each of the 16 COVID patients, we found that Ab-recognition of the RBD of B.1.1.7 variant was quantitatively unaltered, while recognition of the B.1.351 RBD was reduced by approximately 30% (p < 0.0001), and recognition of the P.1 RBD was 50% lower than that for the WT (p < 0.0001) (**Figure 4B**). The reactivity pattern SARS-CoV-2 (WT, D614G) = B1.1.7 > B1.351 > P.1 was seen in each of the COVID-19 patients analyzed.

**Figure 4.**
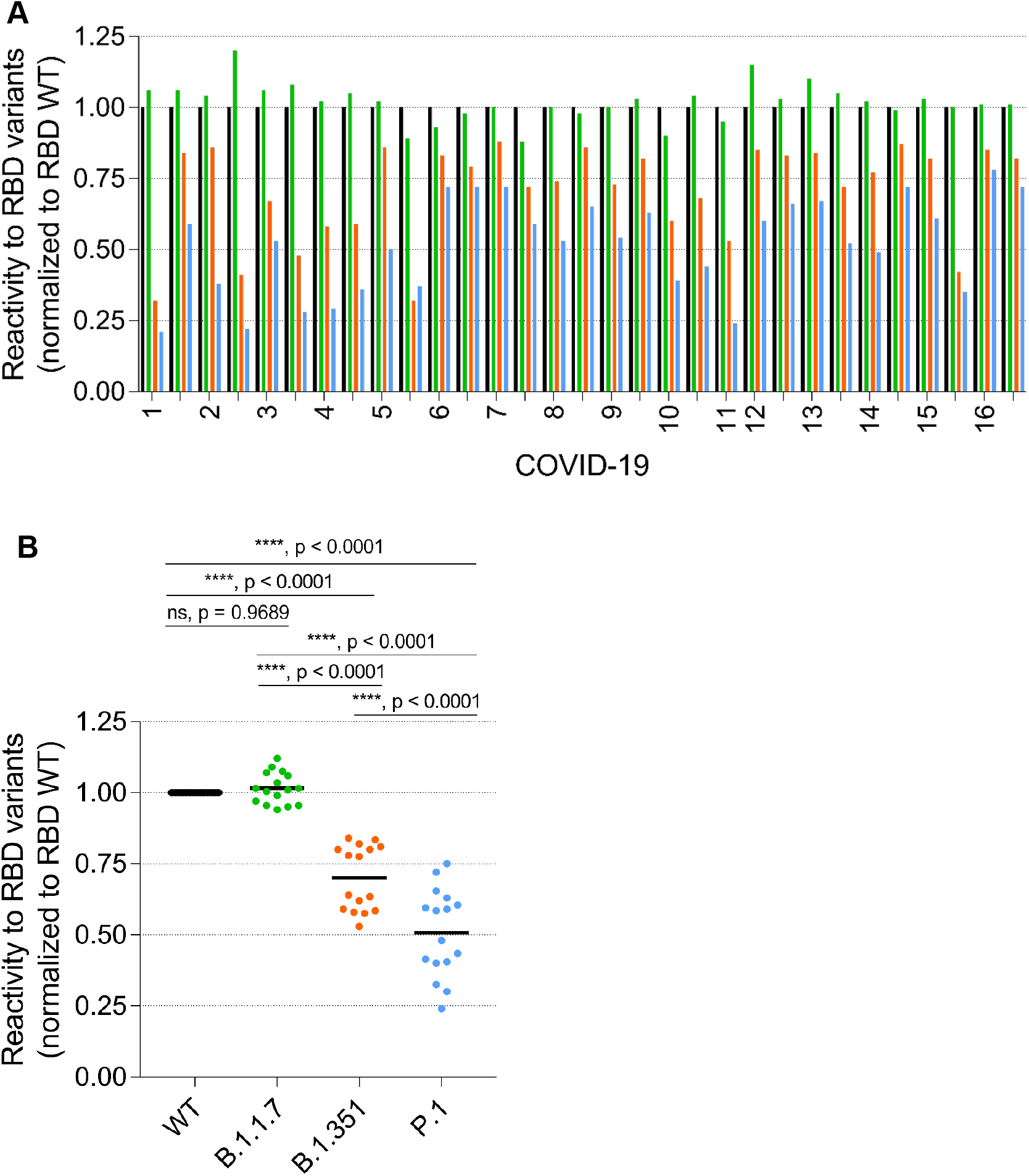
B cell reactivity to RBDs of emerging variants. PBMCs from the indicated COVID-19 patients were differentiated into Ab-secreting cells, and the cell culture supernatants were added to ELISA plates coated with the RBDs of wild-type (WT, black), and of VoCs B.1.1.7/UK (green), B.1.351/SA (orange), or the P.1 lineage, also called B.1.1.248/BR (blue) SARS-CoV-2 variant. (**A**) From #11, one cell culture supernatant was analyzed, from all others two different cell culture supernatants were examined. Reactivity to the wild-type was determined as the delta OD (RBD – BSA) and set as 1, and the relative reactivity to the other RBD variants was calculated and is shown. (**B**) The mean reactivity of the tested wells from each donor was determined. Horizontal bars indicate the mean reactivity to the respective RBD variant (one-way ANOVA, Tukey’s multiple comparison test; each n=16).

## Discussion

In this study, we robustly detected the persistence of memory B cells specific for SARS-CoV-2 in all COVID-19 patients in our cohort that had undergone mild or asymptomatic acute infections, even if their specific serum IgG had declined to undetectable levels (**Figure 5**). This has two implications: Firstly, the persistence of specific memory B cells, which gave rise to neutralizing Abs and showed differential cross-reactivity to VoCs, helps us to understand and start to develop models for predicting long-term protection against SARS-CoV-2. Secondly, the assay employed to differentiate B cells into Ab-producing cells in vitro is a more sensitive method for detecting previous infection than measuring serum levels of SARS-CoV-2 IgG.

**Figure 5.**
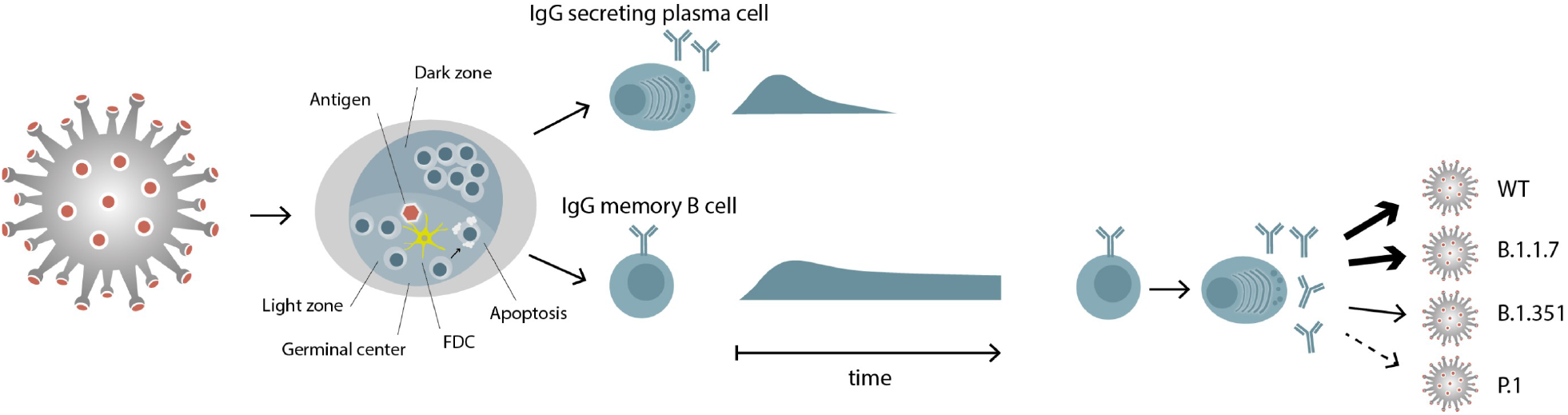
Differential persistence of IgG plasma cells and memory B cells specific for SARS-CoV-2. SARS-CoV-2 antigen is transported to the lymph nodes, where it induces a germinal center reaction with the activation and differentiation of specific B cells. Two types of immune cells exit the germinal center, IgG secreting plasma cells and IgG memory B cells. IgG memory B cells persist, even when the corresponding plasma cells have disappeared. These memory B cells can differentiate to IgG secreting plasma cells and the IgGs they produce show a differential recognition of RBDs of viral variants.

We demonstrated the persistence of specific IgG memory B cells by differentiating them into Ab-secreting cells in vitro, which also facilitated our functional analysis of the Abs secreted by the cells. Previous studies have also detected COVID-19 patients’ memory B cells by staining them with labelled antigens (4, 15, 21, 35-39) and using the enzyme-linked immune absorbent spot (ELISPOT) assay (8, 40-42). Sorting the SARS-CoV-2-specific B cells and cloning their antigen-receptors provided important insights into clonal turnover and the ongoing somatic hypermutation of SARS-CoV-2-specific B cells associated with antigen persistence (15, 21).

We also detected SARS-CoV-2 IgA memory B cells in the blood of the recovered COVID-19 patients, but in contrast to the specific circulating IgG memory B cells, these were only seen in a subset of the cohort and less abundant. While IgA is best known for its role in the immune response at mucosal sites (43, 44), a systemic IgA immune response also occurs that includes the expansion of circulating IgA plasmablasts specific for SARS-CoV-2 (43). Mucosal IgA secretion continues for longer than the serum IgA response (43), but was completely lost after 189 days (43). Circulating IgA declined more rapidly than IgG and decayed by ∼90 days in most COVID-19 cases to levels indistinguishable from controls (4). We robustly detected circulating IgG memory B cells more than 6 months after infection. Taken together, the picture is emerging from this study and previous work that when circulating IgG and IgA, mucosal IgA, and circulating IgA memory B cells are gone, circulating IgG memory B cells persist (**Figure 5**). We found that the Abs produced by these memory B cells had high neutralizing activity, indicating their functional importance upon exposure to the same or mutated SARS-CoV-2.

In general, persisting IgG memory B cells can rapidly differentiate into antibody-secreting cells upon re-exposure (14, 19). The importance of memory B cells for protection against re-infection is evident from immune responses to other human viruses: protective immunity against hepatitis B was observed despite the loss of Abs, (45) and circulating memory B cells to viruses can be sustained for many decades after exposure, well into the tenth decade of life (46). In a mouse model of cytomegalovirus, the activation of virus-specific memory B cells to secrete IgG was independent on cognate or bystander T cell help (47). Thus, the persisting memory B cells are expected to be capable of providing relevant functional protection upon subsequent re-exposure with SARS-CoV-2, even if the relevant Abs have vanished.

The global spread of VoCs such as B.1.1.7, B.1.351, and P.1 may increase re-infection rates and compromise the success of current vaccines (48). We found that the polyclonal memory B cells recognized the RBD of B.1.1.7 (UK variant) to a similar degree as they did the WT, while reactivity to B.1.351 (SA variant) was reduced by 30% and to P.1 (BR variant) by as much as 50%. These findings are in accordance with the molecular signatures of these VoCs, as the UK variant contains one mutation, and the SA and BR variants have three mutations in the RBD (28). A recent study reported a more pronounced drop in cross-reactivity following infection with B.1.1.7 than after infection with the WT, indicating asymmetric heterotypic immunity is induced by SARS-CoV-2 variants (49). Another study showed that some monoclonal (m)Abs showed greatly reduced binding to the UK variant; however, patient serum showed little change in reactivity (50) and unaltered neutralizing activity to the UK variant, but reduced reactivity to the SA variant was reported (51). This is exactly in line with our observations of memory B cells from our cohort. We extended our analysis to show that cross-reactivity for the BR variant was even lower than for the SA variant, which agrees with a recent observation that B.1.351/SA and P.1/BR can escape neutralization by mAbs and are less efficiently inhibited by sera from BNT162b2-vaccinated individuals (28). To what extent the reduced humoral activity is associated with an increased susceptibility to these strains is not yet clear. Re-infection with SARS-CoV-2 is apparently rare on a global scale (52) but was a possible reason for the resurgence of COVID-19 in Manaus (53). It remains to be analyzed whether patients become re-infected because they have lost a certain type of anti-SARS-CoV-2 immunity or are confronted with a new variant, against which they do not yet have a protective immunity.

Because the consequences of infection with SARS-CoV-2 range from asymptomatic to lethal, accurate confirmation of previous infections is of great epidemiological and prognostic significance. Serological testing has made great advances (54-57), but the specific IgG response wanes over time, and some donors with previous infections score negative in current serology tests (4, 16). We showed that the donors who lost IgG to SARS-CoV-2 still had specific IgG memory B cells. It has been previously noted that about 5% of infected donors are “non-responders” who remain seronegative (58, 59). Using our method, we can analyze whether such donors have memory B cells with potential protective activity.

Our assay, involving the differentiation of B cells into Ab-secreting cells in vitro, identifies circulating memory B cells. We distributed the blood cells into different cell-culture-plate wells and analyzed the wells individually to perform an assay with high sensitivity that allows the identification of even rare autoreactive B cells (33). By considering all wells containing samples from each donor, a clear distinction can be made between rare and abundant responses. We noted that, for one HC, one well showed ELISA reactivity to SARS-CoV-2 without neutralizing activity; as the blood sample was obtained before the pandemic, we can probably exclude a previous infection with SARS-CoV-2. The cross-reactivity of memory IgG B cells to SARS-CoV-2, which was detected in only 1/35 wells for all HCs, coincides with previous work showing pre-existing humoral immunity to SARS-CoV-2 in humans (60). However, in our assay, these cross-reactive antibodies did not show neutralizing activity. Nevertheless, such cross-reactivity could be clinically relevant because a recent infection with endemic coronavirus is associated with less severe COVID-19 (61). Thus, the assay we present here extends our methodological armamentarium to evaluate if seronegative individuals have already been infected with SARS-CoV-2. This is of relevance in epidemiology and for optimizing urgently needed immunosuppressive treatments.

Our study had some limitations. The observation of patients was performed up to 8 months after infection. Longer observation times are necessary to learn more about features of B cell persistence. Two donors, who lost SARS-CoV-2-specific IgG but maintained specific memory B cells received immunosuppressive treatment (canakinumab plus hydroxychloroquine or dimethyfumarate) at the time of blood sampling for this study. The impact of immunomodulatory therapies on the maintenance of IgG and memory B cells after infection or vaccination has yet to be analyzed in detail. We analyzed the persistence of B cells reactive to the S1 protein, but the maintenance of B cells against other SARS-CoV-2 proteins remains to be analyzed; although the response to the RBD is of paramount importance, as the RBD is targeted by neutralizing Abs (3). We analyzed circulating memory B cells but are aware that both systemic and mucosal immunity are relevant for protection. We did not analyze the persistence of B cells in patients very severely affected by COVID-19, but it has been reported that the serum Ig response is even greater and longer-lasting in these patients (6, 13).

In this study, we showed that circulating IgG memory B cells specific for SARS-CoV-2 persist in the blood after infection despite the loss of systemic IgG. These persisting B cells can be harnessed to identify a previous infection. Furthermore, these B cells gave rise to neutralizing Abs and showed high cross-reactivity against the emerging UK variant, suggesting patients have protection against re-exposure with this variant even after the loss of specific Abs. In contrast to reactivity against the UK variant, the reactivity of memory B cells against the SA variant and, particularly, the BR variant was greatly reduced. This warrants further attention and indicates a possible need for follow-up vaccinations covering these mutants (62). Thus, our study has added to the efforts to fully uncover the features of SARS-CoV-2-specific memory B cells, which will help us to understand and promote long-term protection.

## Data Availability

Data are available upon request.

## Acknowledgments

This work was supported by the DFG (SFB TR128) and the MOMENTE program LMU (to SM). We are grateful to Prof. M. Kerschensteiner for continuous support and Dipl.–Ing. Benjamin Obholzer for image design. The authors want to thank M.Sc. Samantha Ho and Drs. Lisa Gerdes, Reinhard Hohlfeld, Hartmut Wekerle, and Naoto Kawakami for their comments on the manuscript.

## Author contributions

SW and EM designed the study and experiments. SW, DT, HR, and CS conducted experiments and analyzed data. PE provided reagents and analyzed data. KE, OTK, MK and TK contributed patients’ samples, analyzed data and edited the manuscript. SW, SM, and EM analyzed data and wrote the manuscript.

## Methods

### Study participants

We analyzed the B-cellular responses to SARS-COV-2 of 16 COVID-19-patients (HC=11, MS=4 and SLE=1) that had a mild or asymptomatic disease course (**Table 1**). Five of them were identified in a survey of health care workers (63). Infection was confirmed by SARS-CoV-2-specific PCR (64) and/or SARS-CoV-2 serology, as indicated in **Table 1**. Three patients did not receive a PCR test at the beginning of the pandemic but displayed typical symptoms (fever, respiratory symptoms, severe anosmia) and subsequently tested positive for SARS-CoV-2 Abs. All study participants tested positive for SARS-CoV-2 antibodies at least once after they were infected. As a reference, we analyzed pre-pandemic blood samples from six healthy adults (HC), two males and four females, with a mean age of 28 years.

### Differentiation of B cells into Ab-secreting cells

Our experimental scheme is outlined in **Figure 1**. PBMCs were obtained by a standard density gradient method using SepMate-50 tubes (STEMCELL Technologies, Vancouver, Canada) and frozen. After thawing, the PBMCs were seeded at 1 × 10^6^ cells into 1 mL of culture medium (RPMI + 10 % FCS) in 24-well plates and differentiated into Ab-secreting cells using the TLR7/8 ligand resiquimod (2.5 μg/mL; Sigma-Aldrich, St. Louis, MO, USA) and IL-2 (1,000 IU/mL; R&D Systems, Minneapolis, MN, USA) during 11 days of culturing, essentially as previously described (31-33). This culture system differentiated the memory B cells into Ab-secreting cells (31). The total IgG and IgA levels of the culture supernatants were measured using human IgG and IgA ELISA development kits (Mabtech, Nacka Strand, Sweden). We considered an IgG concentration of > 1 µg/mL to indicate successful differentiation of the B cells and included these wells in the subsequent analysis. To determine the frequency of SARS-CoV-2-specific B cells, 200 μL of PBMCs were distributed as limiting dilutions of between 6.25 × 10^3^ and 6.25 × 10^5^ cells/well in 96-round-bottomed-well plates and stimulated for 11 days culture as described above. The reactivity of the Abs in the supernatants was determined using an RBD ELISA as described below. The antigen-reactive cell frequency was determined according to Poisson distribution as the seeded PBMC count for which 37% of the cultures were negative (31-33). The total B-cell frequency was measured by flow cytometry using the anti-human CD19-PerCP-Cy5.5 Ab (SJ25C1; eBioscience, San Diego, CA, USA), and analysis was performed using FlowJo (10.7.1, BD, Franklin Lakes, NJ, USA).

### ELISAs to detect specific responses to SARS-COV-2 in serum and cell culture supernatants

SARS-CoV-2-specific IgG and IgA were detected in sera and cell culture supernatants using EUROIMMUN anti-SARS-CoV-2 ELISA kits coated with the S1 domain of the SARS-CoV-2 spike protein (EUROIMMUN, Lübeck, Germany). Serum was diluted 1:101 in sample buffer as indicated in the manufacturer’s protocol, and cell culture supernatant was applied undiluted. EUROIMMUN uses a ratio-based analysis for test evaluation and recommends interpreting the results as follows: negative (ratio < 0.8), borderline (ratio ≥ 0.8 to < 1.1), positive (ratio ≥ 1.1). For limiting dilution experiments, Abs against SARS-CoV-2 in the cell culture supernatants were detected using an in-house RBD ELISA. Half-area ELISA plates were coated with 50 µL of RBD (2 μg/mL; P2020-001, Trenzyme, Konstanz, Germany) or bovine serum albumin (BSA, 2 μg/mL; Sigma-Aldrich) overnight at 4°C. The plates were then blocked for 2 h at 37°C with 100 µL of blocking buffer (3% milk in PBS containing 0.05% Tween 20). Undiluted cell culture supernatants (50 µL) were incubated at room temperature for 2 h. Abs were then detected with 50 µL of anti-human IgG horseradish peroxidase (1:5000, 109-036-003, Jackson ImmunoResearch, West Grove, PA, USA) and 50 µL of tetramethylbenzidin (TMB, Sigma-Aldrich) as the substrate. The reaction was stopped by adding 25 µL of 1 M sulfuric acid. The optical density (OD) of the chromogenic reaction was measured at 450 nm, and the plate background was measured at 540 nm. The OD cutoff value for the recognition of RBD was 0.52, which was calculated using the mean + 3 SD of the control cell culture supernatants from stimulated wells with 10^6^ PBMCs per 1 ml.

### Inhibitory activity of Abs towards SARS-CoV-2

To assess the SARS-CoV-2 inhibitory and neutralizing activity of the Abs, we analyzed whether they blocked the binding of the RBD to ACE-2, the cellular receptor for SARS-CoV-2 according to (34) (**Figure 1**). ELISA plates were coated with RBD and blocked as described above. Then 50 µL of the undiluted cell culture supernatants were added and maintained for 2 h at room temperature, followed by the addition of 50 µL of biotin-tagged ACE-2 (1 µg/mL, SAE0171, Sigma-Aldrich). The binding of ACE-2 was detected by 50 µL of streptavidin conjugated with horseradish peroxidase (1:200, 890803, R&D Systems). ELISAs were developed with TMB as described above, and OD values were normalized to those ACE-2 without cell culture supernatant.

### Cross-reactivity to RBD of variants of concern

The cross-reactivities of Abs recognizing SARS-CoV-2 wild type (WT) to RBDs of VoCs B.1.1.7/UK, B.1.351/SA and the P.1 lineage, also called B.1.1.248/BR (28), were determined by ELISA. Half-area ELISA plates were coated with 50 µL of RBD WT (2 μg/mL; PX-COV-P046, ProteoGenix, Schiltigheim, France), RBD B.1.1.7/UK (2 μg/mL; PX-COV-P052, ProteoGenix), RBD B.1.351/SA (2 μg/mL; PX-COV-P053, ProteoGenix), RBD Lineage P.1 also called B.1.1.248/BR (2 μg/mL; PX-COV-P054, ProteoGenix), or BSA (2 μg/mL; Sigma-Aldrich) overnight at 4°C. The subsequent procedure was as described for the RBD ELISA.

### Statistics

Statistical analyses were performed using GraphPad Prism 7 (GraphPad Software Inc., La Jolla, CA, USA).

### Study approval

The study was approved by the ethical committee of the medical faculty of the Ludwig-Maximilians-Universität Munich. Written informed consent was obtained from each donor prior to their inclusion in the study.

