## Supplemental Figure 1 for "Persistence of functional memory B cells recognizing SARS-CoV-2 variants despite loss of specific IgG"

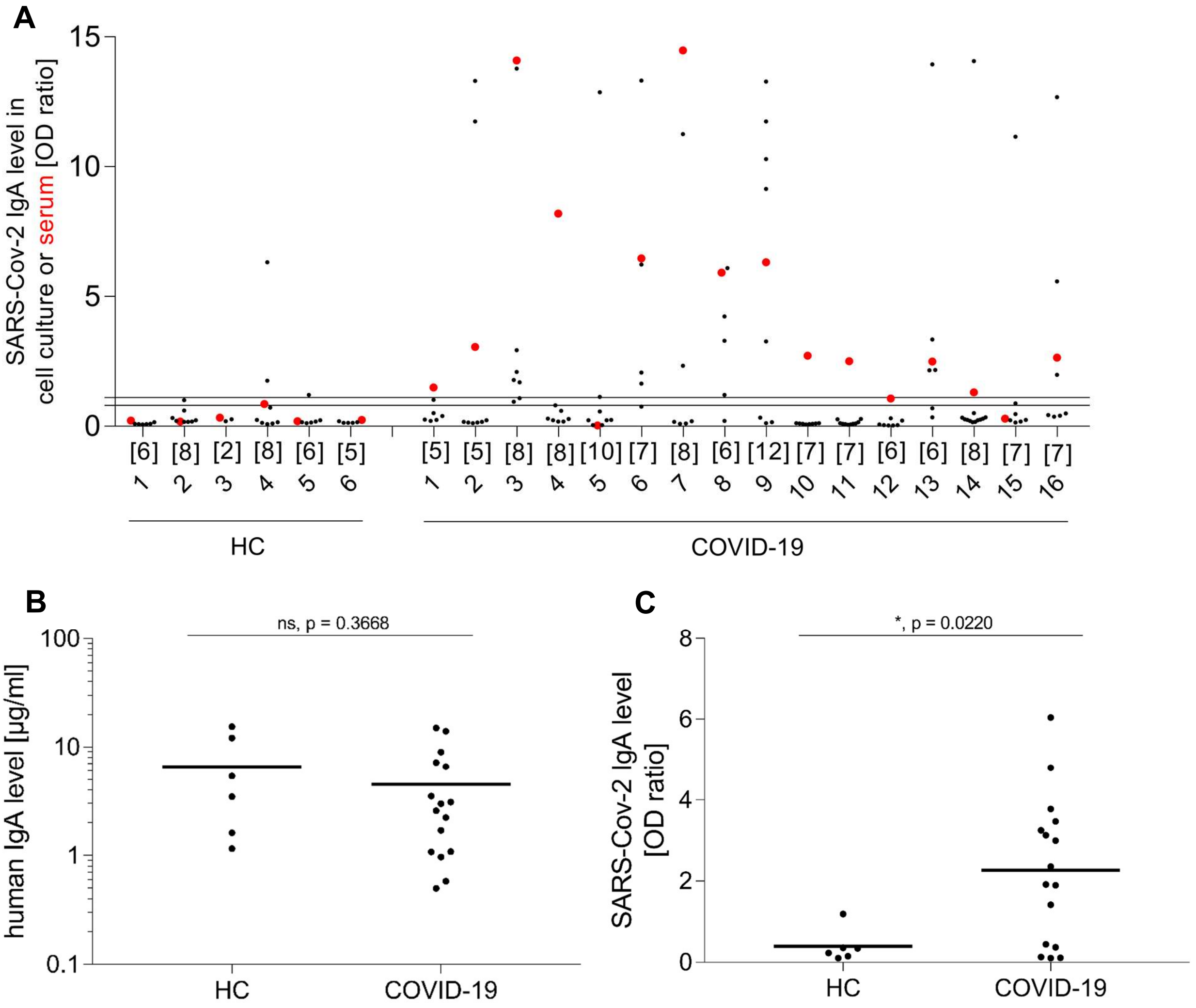


**Supplemental Figure 1. Systemic IgA response to SARS-CoV-2**

(**A**) PBMCs from heathy controls (HC, left) and COVID-19-patients (right) were differentiated into Ab-secreting cells. The reactivity of IgA in cell culture supernatants against S1 was determined. Each black dot represents one stimulated well. The number of stimulated wells per donor is provided directly under the x-axis. The reactivity of the serum in the same ELISA is shown with a red dot. The area between the two horizontal lines was considered the borderline zone. (**B, C**) Each symbol represents the mean IgA levels of all stimulated wells in one donor. Horizontal lines indicate the mean IgA levels of all donors in the respective groups. (**B**) IgA levels of cell culture supernatants were not significantly different between the groups ($p = 0.3668$, Mann-Whitney U; HC=6, COVID-19=16). (**C**) The COVID-19 group had more anti-S1 IgA than HC ($p < 0.05$, Mann-Whitney U; HC=6, COVID-19=16).
